# Measuring circadian rhythms in patients with dementia: protocol for a scoping review

**DOI:** 10.1101/2022.06.06.22275886

**Authors:** Victoria G. Gabb, Louise M. Ince, Sarah Rudd, Elizabeth Coulthard

## Abstract

**Introduction:** Most physiological processes are under circadian regulation. Disturbances to circadian rhythms are common and often early symptoms in dementia. There is a gap in the literature regarding how and which circadian functions have been, or could optimally be, measured in patients with dementia. This protocol outlines a scoping review to identify which and how circadian variables have been studied in patients with dementia and mild cognitive impairment, and examine whether feasibility, acceptability, appropriateness, and utility in clinical practice has been considered.

**Methods and analysis:** The scoping review will be conducted in accordance with the Joanna Briggs Institute methodology for scoping reviews and will be reported in line with the Preferred Reporting Items for Systematic reviews and Meta-Analyses extension for Scoping Reviews (PRISMA-ScR). We will systematically search five electronic bibliographic databases (MEDLINE, CINAHL, EMBASE, The Cochrane Central Register of Controlled Trials (CENTRAL), PsycInfo). There will be no restriction on date of publication or language; corresponding authors will be contacted where relevant non-English articles are identified to request a translation. Search results will be merged using reference management software and duplicates will be removed. To ensure consistency in applying eligibility criteria, 10% of retrieved articles will be checked by each reviewer in the team at the title and abstract screening stage and full texts will be reviewed by at least 2 reviewers. Data from eligible articles will be extracted using a standardised form. Where study outcomes (e.g., patient and public involvement, adherence) have not been described in the article, authors will be contacted in case relevant data is available to share but was omitted from the main report. Results will be presented as a narrative synthesis as study and population heterogeneity will likely prohibit meta-analysis.

**Ethics and dissemination:** Ethical approval is not required as the scoping review methodology does not involve human or animal participants. This review will provide an extensive overview of established and emerging trends in measurement of circadian rhythms in dementia and identify gaps in the research to inform future clinical and research practice. The review will also highlight whether patient and public involvement, participant burden, and clinical usefulness of methods have been considered in developing protocols. Results will be disseminated in a peer-reviewed publication.

## Introduction

Often considered in relation to the sleep-wake cycle, circadian rhythms are a prominent feature of human biology observed across many physiological parameters including cell division, hormone secretion, metabolism, temperature regulation, and behavioural rhythms ^(1, 2)^. Circadian rhythms, approximately 24-hour cycles coordinated to the light-dark cycle of the Earth, are generated on a cellular level by molecular clocks and synchronised by humoral and neural signals coordinated by the brain’s central pacemaker, the suprachiasmatic nuclei ^(1,3)^.

Increasing age is associated with significant changes in circadian rhythms ^(4-7)^. Many rhythmic parameters investigated in healthy older adults exhibit dampened oscillations, whilst others undergo ‘phase advance’ with rhythms shifting to earlier in the day ^(8,9)^. Sleep-wake cycles become disrupted ^(10,11)^, and rhythms in many circulating hormone concentrations become flattened ^(12-14)^. Some of these changes are considered inevitable changes associated with physiological ageing, whilst others may be pathological signals due to the vulnerability to neurodegeneration with increasing age ^(15-19)^.

Circadian disruptions are frequently exacerbated in populations with cognitive decline and dementia ^(11, 18-20)^. Monitoring the rate of circadian rhythm breakdown may be a clinically useful tool to assess progression of underlying pathology and strategies to improve circadian function may represent a useful therapeutic strategy. Reviews have begun to emerge exploring changes in circadian functions in this population, focusing on causative mechanisms and initial findings ^(15-18, 21)^. However, there is scarce research into the practical or ethical considerations of measuring circadian function in adults with dementia. Traditional methods of measuring circadian dysfunction, including forced desynchrony protocols, sleep deprivation, and repetitive blood tests are burdensome for patients and may exacerbate distressing symptoms, while less rigorous protocols may compromise data integrity. Most reviews to date have also focused on Alzheimer’s disease and sleep-wake cycles, without covering related neurodegenerative dementias, such as vascular or Lewy body dementias, or other circadian rhythms ^(22-25)^.

This scoping review will evaluate two methodological aspects of circadian research in adults with dementia and mild cognitive impairment (MCI). Firstly, the review aims to examine which circadian functions have been reported and how these outcomes have been measured. Secondly, the review aims to provide an overview of the extent to which clinicians, patients, or the public have been consulted on acceptability of technique in research and/or clinical use. A review of existing research methods, including consideration of their benefits and drawbacks and gaps in our knowledge, will provide recommendations for getting the most benefit from future research into circadian function in dementia.

### Review question

“How have circadian rhythms been measured in research involving participants with dementia or MCI?”

Specifically, we will review:

- Which physiological processes have been examined in patients with dementia/MCI in relation to circadian functioning?
- What techniques and protocols have been used to study circadian rhythms and have these been validated in the population?
- Considering quantitative data (e.g., adherence, attrition rates), qualitative data (e.g., feedback from participants and other stakeholders), participant burden, and invasiveness, were the methods used to study circadian function feasible and acceptable in patients with dementia/MCI?
- What key learning has been reported with regards to measuring circadian rhythms in patients with dementia/MCI and how could this learning feed into recommendations for best practice in future research?

### Eligibility criteria

#### Participants

We will include studies with human adult participants (aged 18 and over) who have a diagnosis of neurodegenerative dementia or mild cognitive impairment (MCI) where diagnosis meets established diagnostic criteria (e.g., Albert criteria, DSM-5, ICD-11).

#### Concept

To be eligible for inclusion, the study must examine the circadian rhythm of at least one physiological process as an outcome of interest.

Studies which investigate related behaviours (e.g., sleep or eating patterns) without an explicit focus on circadian rhythm will be excluded. Studies which attempt to experimentally manipulate circadian function will only be included if they also measure a circadian outcome (e.g. a study examining bright light therapy which measures melatonin will be eligible for inclusion, whilst a study administering melatonin treatment but not measuring a circadian outcome will be ineligible).

#### Context

The review will include studies conducted in any setting.

#### Types of evidence sources

This scoping review will consider quantitative and qualitative primary research studies, including experimental, quasi-experimental, and observational studies, including but not limited to: randomized and non-randomized controlled trials, interrupted time-series studies, prospective and retrospective cohort studies, case-control studies, and cross-sectional studies. Protocols, trial registry details, and unpublished (grey) literature such as conference abstracts and theses will be included where published articles for the same study are unavailable, or where they provide additional information to answer the review question alongside a published article. Reviews, text, and opinion papers will not be eligible for inclusion in the main scoping review as they constitute secondary data sources.

### Methods

The scoping review will be conducted in accordance with the JBI methodology for scoping reviews ^(26)^ and will be reported in line with the Preferred Reporting Items for Systematic reviews and Meta-Analyses extension for Scoping Reviews (PRISMA-ScR) ^(27)^.

### Search strategy

The search strategy will aim to locate both published and unpublished studies ^(26)^. An initial scoping search of MEDLINE and CINAHL was undertaken to identify key words contained in the titles and abstracts and Medical Subject Headings (MeSH) index terms. In conjunction with a medical librarian, a full search strategy will be developed using relevant text words and subject headings for “dementia” and “circadian rhythms” and translated for the following databases: EMBASE (Ovid), Medline (Ovid), CINAHL (Ebsco), PsycInfo (ProQuest) and Cochrane Central Register of Controlled Trials (CENTRAL).

The reference list of all sources of evidence identified as potentially eligible at the title and abstract screening stage will be screened for additional relevant studies.

No restriction on publication date will be applied to identify any trends over time in measurement of circadian function in this population.

Studies available in English will be included; authors will be contacted to request translations for studies published in other languages. Authors of all included studies will be contacted for further information on their experience of researching circadian function in patients with dementia/MCI, including recruitment rates, adherence, feedback from participants, and engagement in patient and public involvement (PPI) activities during the study period. Where searches identify unpublished (grey) literature, protocols, or trial registry entries, the reviewer will seek to identify the same study in published literature. If unavailable, the unpublished literature will be included in the study.

### Study selection

Following the search, all identified citations will be collated and uploaded into reference management software and duplicates removed. Titles and abstracts will be independently screened by two or more reviewers for assessment against the inclusion criteria for the review. Full texts will be reviewed in detail against the inclusion criteria by two independent reviewers with discrepancies resolved via discussion or involvement of a third reviewer.

Reasons for exclusion at the full-text review stage will be recorded and reported in the scoping review. Any disagreements that arise between the reviewers at each stage of the selection process will be resolved through discussion. The results of the search and the study inclusion process will be reported in full in the final scoping review using an adapted Preferred Reporting Items for Systematic Reviews and Meta-analyses (PRISMA) flow diagram template ^(28)^ to report the number of search results generated, the number of articles excluded at each stage, and the final number of included studies.

### Data extraction

Data will be extracted from eligible full-text papers by two or more independent reviewers using a data extraction tool developed by the reviewers. The data extracted will include specific details about the study characteristics (e.g., year, country, study aims and design), participants (e.g., diagnosis, age, sex, ethnicity), and study methods and key outcomes relevant to the review question (e.g., sampling protocol and method, circadian proxy/variables of interest, feedback from patient groups). The data extraction tool will be revised as necessary during the process of extracting data from each included article following discussion with the reviewing team. The final data extraction form will be included in the scoping review.

### Data analysis and presentation

Study characteristics of included articles will be summarised in a table and a brief narrative overview will be provided. The results section will answer how circadian function has been measured in individuals with dementia/MCI, focusing predominantly on sampling techniques and protocols and evidence of acceptability, feasibility, and appropriateness of the methods used. The report will involve narrative syntheses complemented by frequency tables. Graphs will be used to illustrate salient findings.

## Data Availability

No data is presented in this manuscript.

## Acknowledgements

VGG and LMI wrote the original draft and developed the extraction form and initial search strategy. SR substantively revised and finalised the search strategy. All authors have contributed to the manuscript and read and approved the final version.

